# Spatial and temporal regularization to estimate COVID-19 Reproduction Number *R*(*t*): Promoting piecewise smoothness via convex optimization

**DOI:** 10.1101/2020.06.10.20127365

**Authors:** Patrice Abry, Nelly Pustelnik, Stéphane Roux, Pablo Jensen, Patrick Flandrin, Rémi Gribonval, Charles-Gérard Lucas, Eric Guichard, Pierre Borgnat, Nicolas Garnier, Benjamin Audit

## Abstract

Among the different indicators that quantify the spread of an epidemic, such as the on-going COVID-19, stands first the reproduction number which measures how many people can be contaminated by an infected person. In order to permit the monitoring of the evolution of this number, a new estimation procedure is proposed here, assuming a well-accepted model for current incidence data, based on past observations. The novelty of the proposed approach is twofold: 1) the estimation of the reproduction number is achieved by convex optimization within a proximal-based inverse problem formulation, with constraints aimed at promoting piecewise smoothness; 2) the approach is developed in a multivariate setting, allowing for the simultaneous handling of multiple time series attached to different geographical regions, together with a spatial (graph-based) regularization of their evolutions in time. The effective-ness of the approach is first supported by simulations, and two main applications to real COVID-19 data are then discussed. The first one refers to the comparative evolution of the reproduction number for a number of countries, while the second one focuses on French counties and their joint analysis, leading to dynamic maps revealing the temporal co-evolution of their reproduction numbers.

## 1 Introduction

### Context

The ongoing COVID-19 pandemic has produced an unprecedented health and economic crisis, urging for the development of adapted actions aimed at monitoring the spread of the new coronavirus. No country remained untouched, and all of them experienced a propagation mechanism that is basically universal in the onset phase: each infected person happened to infect in average more than one other person, leading to an initial exponential growth.

The strength of the spread is quantified by the so-called *reproduction number* which measures how many people can be contaminated by an infected person. In the early phase where the growth is exponential, this is referred to as *R*_0_ (for COVID-19, *R*_0_ ∼ 3 [13, 23]). As the pandemic develops and because more people get infected, the effective reproduction number evolves, hence becoming a function of time hereafter labeled *R*(*t*). This can indeed end up with the extinction of the pandemic, *R*(*t*) → 0, at the expense though of the contamination of a very large percentage of the total population, and of potentially dramatic consequences.

Rather than letting the pandemic develop until the reproduction number would eventually decrease below unity (in which case the spread would cease by itself), an active strategy amounts to take actions so as to limit contacts between individuals. This path has been followed by several countries which adopted effective *lock-down* policies, with the consequence that the reproduction number decreased significantly and rapidly, further remaining below unity as long as social distancing measures were enforced (see for example [14, 23]).

However, when lifting the lock-down is at stake, the situation may change with an expected increase in the number of inter-individual contacts, and monitoring in real time the evolution of the instantaneous reproduction number *R*(*t*) becomes of the utmost importance: this is the core of the present work.

### Issues and related work

Monitoring and estimating *R*(*t*) raises however a series of issues related to pandemic data modeling, to parameter estimation techniques and to data availability. Concerning the mathematical modeling of infectious diseases, the most celebrated approaches refer to *compartmental models* such as SIR (“Susceptible - Infectious - Recovered”), with variants such as SEIR (“Susceptible - Exposed - Infectious - Recovered”). Because such global models do not account well for spatial heterogeneity, clustering of human contact patterns, variability in typical number of contacts (cf. [15]), further refinements were proposed [3]. In such frameworks, the effective reproduction number at time *t* can be inferred from a fit of the model to the data that leads to an estimated knowledge of the average of infecting contacts per unit time, of the mean infectious period, and of the fraction of the population that is still susceptible. These are powerful approaches that are descriptive and potentially predictive, yet at the expense of being fully parametric and thus requiring the use of dedicated and robust estimation procedures. Parameter estimation become all the more involved when the number of parameters grows and/or when the amount and quality of available data are low, as is the case for the COVID-19 pandemic *real-time* and *in emergency* monitoring.

Rather than resorting to fully parametric models and seeing *R*(*t*) as the by-product of its identification, a more phenomenological, semi-parametric approach can be followed [11, 18, 25]. This approach has been reported as robust and potentially leading to relevant estimates of *R*(*t*), even for epidemic spreading on realistic contact networks, where it is not possible to define a steady exponential growth phase and a basic reproduction number [15]. The underlying idea is to model incidence data *z*(*t*) at time *t* as resulting from a Poisson distribution with a time evolving parameter adjusted to account for the data evolution. This parameter can be written as 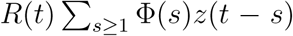, where *z*(*t* −*s*) accounts for the past incidence data, as convolved with a function Φ(*s*) standing for the distribution of the *serial interval*. The serial interval function Φ(*s*) models the time between the onset of symptoms in a primary case and the onset of symptoms in secondary cases, or equivalently the probability that a person confirmed infected today was actually infected *s* days earlier by another infected person. The serial interval function is thus an important ingredient of the model, accounting for the biological mechanisms in the epidemic evolution.

Assuming the distribution Φ to be known (which can be questionable), the whole challenge in the actual use of the semi-parametric Poisson-based model thus consists in devising estimates 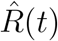 of *R*(*t*) that have better statistical performance (more robust, reliable and hence usable) than the direct brute-force and naive form:

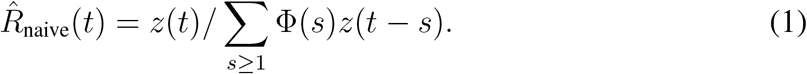

This has been classically addressed by approaches aimed at maximizing the likelihood attached to the model. This can be achieved, e.g., within several variant of Bayesian frameworks [1, 11, 14, 15, 25], yet at the expense of a heavy computational burden. We promote here an alternative approach based on inverse problem formulations and proximal-operator based nonsmooth convex optimisation [2, 4, 9, 19, 21].

The questions of modeling and estimation, be they fully parametric or semi-parametric, are intimately intertwined with that of data availability. This will be discussed in further detail in Section 2, but one can however remark at this point that many options are open, with a conditioning of the results to the choices that are made. There is first the nature of the incidence data used in the analysis (reported infected cases, hospitalizations, deaths) and the database they are extracted from. Next, there is the granularity of the data (whole country, regions, smaller units) and the specificities that can be attached to a specific choice as well as the comparisons that can be envisioned. In this respect, it is worth remarking that most analyses reported in the literature are based on (possibly multiple) univariate time series, whereas genuinely multivariate analyses (e.g., a joint analysis of the same type of data in different countries in order to compare health policies) might prove more informative.

### Goal, contributions and outlines

For that category of research work motivated by contributing *in emergency* to the societal stake of monitoring the pandemic evolution in *realtime*, or at least, on a *daily basis*, there are two classes of challenges: ensuring a robust and regular access to relevant data; rapidly developing analysis/estimation tools that are theoretically sound, practically usable on data actually available, and that may contribute to improving current monitoring strategies. In that spirit, the overarching goal of the present work is twofold: (1) proposing a new, more versatile framework for the estimation of *R*(*t*) within the semi-parametric model of [11, 25], reformulating its estimation as an inverse problem whose functional is minimized by using non smooth proximal-based convex optimization; (2) inserting this approach in an extended multivariate framework, with applications to various, complementary datasets corresponding both to different incidence data and to different geographical regions.

The data used here were collected from three different databases (Johns Hopkins University, European Centre for Disease Prevention and Control, and Santé-Publique-France). While incidence data reported essentially consist of the number of infected, hospitalized, dead and recovered persons, the three databases are very heterogenous with respect to starting date of data availability, geographical granularity, and data quality (outliers, misreporting,…). This is detailed in Section 2.1. In the present work, it has been chosen to work with the number of daily new infections as incidence data, thus labeled *z*(*t*) in the remainder of the work. Data other than the number of confirmed cases are not studied here, for reasons further discussed in Section 5. The number of daily new infections may however be directly *read* as reported in databases, or *recomputed* from other available data (such as hospitalization numbers). Further, the uneven quality of the data is such that preprocessing has proved necessary. These issues are detailed in Section 2.2. Section 3 presents the semi-parametric model for *R*(*t*) (cf. Section 3.1) and how its estimation can be phrased within a non smooth proximal-based convex optimization framework, intentionally designed to enforce piecewise linearity in the estimation of *R*(*t*) via temporal regularization, as well as piecewise constancy in spatial variations of *R*(*t*) by graph-based regularization (cf. Section 3.2). Proximal-operator based algorithms for the minimization for the corresponding functionals are detailed in Section 3.3. These estimation tools have been first illustrated at work on synthetic data, constructed from different models and simulating several scenarii (cf. Section 3.4). They were then applied to several real pandemic datasets (cf. Section 4). First, the number of daily new infections for many different countries across the world were analyzed independently (cf. Section 4.3). Second, focusing on France only, the number of daily new infections per continental France *départements* (*départements* constitute usual entities organizing the administrative life in France) were analyzed both independently and in a multivariate setting, illustrating the benefit of this latter formulation (cf. Section 4.4). Discussions, perpectives and potential improvements are discussed in Section 5.

## 2 Data

### 2.1 Datasets

To enable a relevant study of the pandemic, it is essential to have at disposal robust and automated accesses to reliable databasets where pandemic-related data are made available by relevant authorities, on a regular basis. In the present study, three sources of data were systematically used.

#### Source1(JHU)

Johns Hopkins University^1^ provides access to the cumulated daily reports of the number of infected, deceased and recovered persons, on a per country basis, for a large number of countries worldwide, essentially since inception of the COVID-19 crisis (January 1st, 2020).

#### Source2(ECDPC)

The European Centre for Disease Prevention and Control^2^ provides similar information.

#### Source3(SPF)

Santé-Publique-France^3^ focuses on France only. It makes available on a daily basis a rich variety of pandemic-documented data across the France territory on a per *département*-basis, *départements* consisting of the usual granularity of geographical units (of roughly comparable sizes), used in France to address most administrative issues (an equivalent of counties for other countries). Source3(SPF) data are mostly based on hospital records, such as the daily reports of the number of currently hospitalized persons, together with the cumulated numbers of deceased and recovered persons with breakdowns by age and gender. Elementary algebra enables us to derive the daily number of new hospitalizations, used as a (delayed) proxy for daily new infections, assuming that a constant fraction of infected people is hospitalized. Data are however available only after March 20th.

Data were (and still are) automatically downloaded on a daily basis, using MATLAB routines, written by ourselves and available upon request.

### 2.2 Time series

The data available on the different data repositories used here are strongly affected by outliers, which may stem from inaccuracy or misreporting in per country reporting procedures, or from changes in the way counts are collected, aggregated, and reported. Outliers are present for all countries and all datasets, and occur in related manners. Accounting for outliers is per se an issue that can be handled in several different ways (this is further discussed in Section 5). In the present work, it has been chosen to preprocess data for outlier removal by applying to the raw time series a nonlinear filtering, consisting of a sliding-median over a 7-day window: outliers defined as ±2.5 standard deviation are replaced by window median to yield the pre-processed time series *z*(*t*), from which the reproduction number *R*(*t*) is estimated. Examples of raw and pre-processed time series are illustrated in the figures throughout Section 4.

Countries are studied independently (cf. Section 4.3), and the estimation procedure is thus applied independently to each time series *z*(*t*) of size *T*, the number of days available for analysis.

To each continental France *département* is associated a time series *z*_*d*_(*t*) of size *T*, where 1 ≤*d* ≤ *D* = 94 indexes the *département*. These time series are collected and stacked in a matrix of size *D* × *T*, and are analyzed both independently and jointly (cf. Section 4.4).

Estimation of *R*(*t*) is performed daily, with *T* thus increasing every day.

## 3 Data model and estimation procedure

### 3.1 Data modeling

As mentioned in Introduction, epidemiological data are often modeled by SIR models and variants, devised to account for the detailed mechanisms driving the epidemic outbreak. Although they can be used for envisioning the impact of possible scenarii in the future development of an on-going epidemic [13], such models, because they require the full estimation of numerous parameters, are often used a posteriori (e.g., long after the epidemic) with consolidated and accurate datasets. During the spread phase and in order to account for the on-line/on-the-fly need to monitor the pandemic and to offer some robustness to partial/incomplete/noisy data, less detailed semi-parametric models focusing on the only estimation of the time-dependent reproduction number can be preferred [26, 18, 11].

Let *R*(*t*) denote the instantaneous reproduction number to be estimated and *z*(*t*) be the number of daily new infections. It has been proposed in [11, 25] that {*z*(*t*), *t* = 1, …, *T*} can be modeled as a nonstationary time series consisting of a collection of random variables, each drawn from a Poisson distribution *𝒫*_*p*_*t* whose parameter *p*_*t*_ depends on the past observations of *z*(*t*), on the current value of *R*(*t*), and on the *serial interval* function Φ(·):

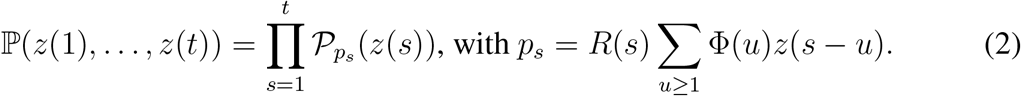

The *serial interval* function Φ(·) constitutes a key ingredient of the model, whose importance and role in pandemic evolution has been mentioned in Introduction. It is assumed to be independent of calendar time (i.e., constant across the epidemic outbreak), and, importantly, independent of *R*(*t*), whose role is to account for the time dependencies in pandemic propagation mechanisms.

For the COVID-19 pandemic, several studies have empirically estimated the serial interval function Φ(·) [16, 22]. For convenience, Φ(·) has been modeled as a Gamma distribution, with shape and rate parameters 1.87 and 0.28, respectively (corresponding to mean and standard deviations of 6.6 and 3.5 days, see [14] and references therein). These choices and assumptions have been followed and used here, and the corresponding function is illustrated in Fig. 1.

**Figure 1.**
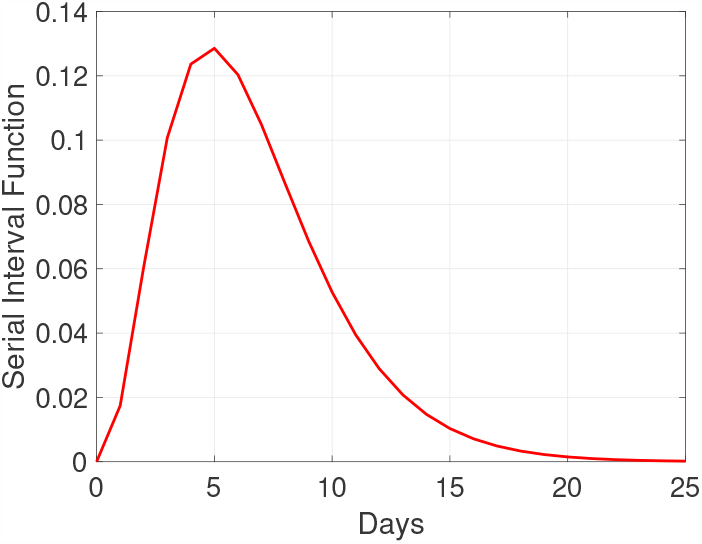
**Serial interval function** Φ modeled as a Gamma distribution with mean and standard deviation of 6.6 and 3.5 days, following [16].

In essence, the model in Eq. (2) is *univariate* (only one time series is modeled at a time), and based on a Poisson marginal distribution. It is also *nonstationary*, as the Poisson rate evolves along time. The key ingredient of this model consists of the Poisson rate evolving as a weighted moving average of past observations, which is qualitatively based on the following rationale: when 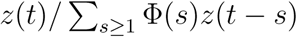 is above 1, the epidemic is growing and, conversely, when this ratio is below 1, it decreases and eventually vanishes.

### 3.2 Estimation via non-smooth convex optimisation

In order to estimate *R*(*t*), and instead of using Bayesian frameworks that are considered state-of-the-art tools for epidemic evolution analysis, we propose and promote here an alternative approach based on an inverse problem formulation. Its main principle is to assume some form of *temporal regularity* in the evolution of *R*(*t*) (below we use a piecewise linear model). In the case of a joint estimation of *R*(*t*) across several continental France *départements*, we further assume some form of *spatial regularity*, i.e., that the values of *R*(*t*) for neighboring *départements* are similar.

#### Univariate setting

For a single country, or a single *département*, the observed (possibly pre-processed) data {*z*(*t*), 1 ≤ t ≤ T} is represented by a *T*-dimensional vector **z** ∈ ℝ^*T*^. Recalling that the Poisson law is 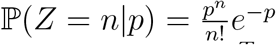 for each integer *n* ≥ 0, the negative log-likelihood of observing **z** given a vector **p** *∈* ℝ^*T*^ of Poisson parameters *p*_*t*_ is

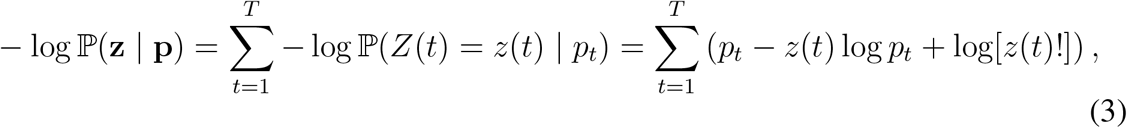

where **r** ∈ ℝ^*T*^ is the (unknown) vector of values of *R*(*t*). Up to an additive term independent of **p**, this is equal to the KL-divergence (cf. Section 5.4. in [21]):

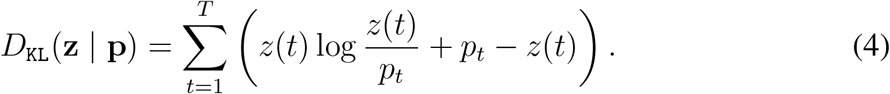

Given the vector of observed values **z**, the serial interval function Φ(*·*), and the number of days *T*, the vector **p** given by (2) reads **p** = **r** *0* **Φz**, with *0* the entrywise product and **Φ** *∈* ℝ^*T ×T*^ the matrix with entries **Φ**_*ij*_ = Φ(*i − j*).

Maximum likelihood estimation of **r** (i.e., minimization of the negative log-likelihood) leads to an optimization problem min_**r**_ *D*_KL_(**z** | **r** 0 **Φz**) which does not ensure any regularity of *R*(*t*). To ensure *temporal* regularity, we propose a penalized approach using 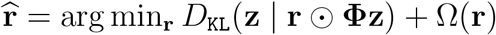 where Ω denotes a penalty function.

Here we wish to promote a piecewise affine and continuous behavior, which may be accomplished [7, 12] using Ω(**r**) = *λ*_time_ ∥**D**_2_**r** ∥_1_, where **D**_2_ is the matrix associated with a Laplacian filter (second order discrete temporal derivatives), ∥ ∥_1_ denotes the */J*_1_-norm (i.e., the sum of the absolute values of all entries), and *λ*_time_ is a penalty factor to be tuned. This leads to the following optimization problem:

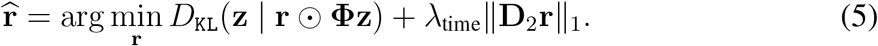

#### Spatially regularized setting

In the case of multiple *départements*, we consider multiple vectors (**z**_*d*_ ∈ ℝ^*T*^, 1 ≤ *d* ≤*D*) associated to the *D* time series, and multiple vectors of unknown (**r**_*d*_ ∈ ℝ^*T*^, ≤1 ≤*d D*), which can be gathered into matrices: a data matrix **Z** ∈ ℝ^*T ×D*^ whose columns are **z**_*d*_ and a matrix of unknown **R** ∈ ℝ^*T ×D*^ whose columns are the quantities to be estimated **r**_*d*_.

A first possibility is to proceed to independent estimation of the (**r**_*d*_ ∈ ℝ^*T*^, ≤1 *d* ≤ *D*) by addressing the separate optimization problems

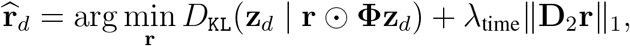

which can be equivalently rewritten into a matrix form:

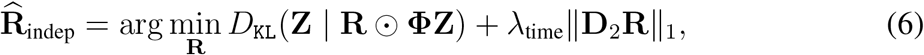

Where 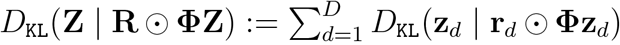, and 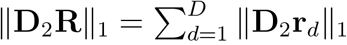 is the entrywise *ℓ*^1^ norm of **D**_2_**R**, i.e., the sum of the absolute values of all its entries.

An alternative is to estimate jointly the (**r**_*d*_ ∈ ℝ^*T*^, ≤1 *d* ≤ *D*) using a penalty function promoting *spatial* regularity. To account for spatial regularity, we use a spatial analogue of **D**_2_ promoting spatially piecewise constant solutions. The *D* continental France *départements* can be considered as the vertices of a graph, where edges are present between adjacent *départements*. From the adjacency matrix **A** ∈ ℝ^*D×D*^ of this graph (**A**_*ij*_ = 1 if there is an edge *e* = (*i, j*) in the graph, **A**_*ij*_ = 0 otherwise), the global variation of the function on the graphs can be computed as ∑_*ij*_ **A**_*ij*_(**R**_*ti*_ **R**_*tj*_)^2^ and it is known that this can be accessed through the so-called (combinatorial) Laplacian of the graph: **L** = Δ *−* **A** where Δ is the diagonal matrix of the degrees (Δ_*ii*_ = ∑_*j*_ **A**_*ij*_) [24]. However, in order to promote smoothness over the graphs while keeping some sparse discontinuities on some edges, it is preferable to regularize using a Total Variation on the graph, which amounts to take the */J*_1_-norm of these gradients (**R**_*ti*_ *−* **R**_*tj*_) on all existing edges. For that, let us introduce the incidence matrix **B** *∈* ℝ^*E×D*^ such that **L** = **B**^T^**B** where *E* is the number of edges and, on each line representing an existing edge *e* = (*i, j*), we set **B**_*e,i*_ = 1 and **B**_*e,j*_ = *−*1. Then, the *ℓ*_1_-norm ∥**RB**^T^ ∥_1_ = ∥**BR**^T^ ∥_1_ is the equal to 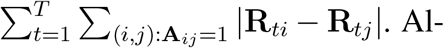 ternatively, it can be computed as 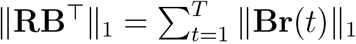 where **r**(*t*) ℛ R^*D*^ is the *t*-th row of **R**, which gathers the values across all *départements* at a given time *t*. From that, we can define the regularized optimization problem:

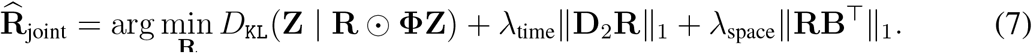

Optimization problems (6) and (7) involve convex, lower semi-continuous, proper and non-negative functions, hence their set of minimizers is non-empty and convex [2]. We will soon discuss how to compute these using proximal algorithms. By the known sparsity-promoting properties of *ℓ*^1^ regularizers and their variants, the corresponding solutions are such that **D**_2_**R** and/or **RB**^*T*^ are sparse matrices, in the sense that these matrices of (second order temporal or first order spatial) derivatives have many zero entries. The higher the penalty factors *λ*_time_ and *λ*_space_, the more zeroes in these matrices. In particular, when *λ*_space_ = 0 no spatial regularization is performed, and (7) is equivalent to (6). When *λ*_space_ is large enough, **RB**^*T*^ is *exactly* zero, which implies that **r**(*t*) is constant at each time since the graph of *départements* is connected. How to tune such parameters is further discussed in Section 4.1.

### 3.3 Optimization using a proximal algorithm

The considered optimization problems are of the form

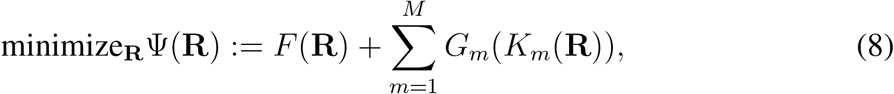

where *F* and *G*_*m*_ are proper lower semi-continuous convex, and *K*_*m*_ are bounded linear operators. A classical case for *m* = 1 is typically addressed with the Chambolle-Pock algorithm [5], which has been recently adapted for multiple regularization terms as in Eq. 8 of [10]. To handle the lack of smoothness of Lipschitz differentiability for the considered functions *F* and *G*_*m*_, these approaches rely on their proximity operators. We recall that the proximity operator of a convex, lower semi-continuous function *φ* is defined as [17]

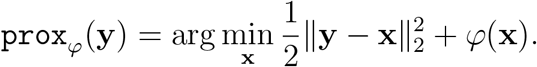

In our case, we consider a separable data fidelity term:

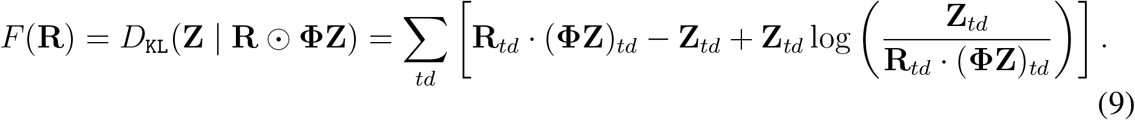

As this is a separable function of the entries of its input, its associated proximity operator can be computed component by component [8]:

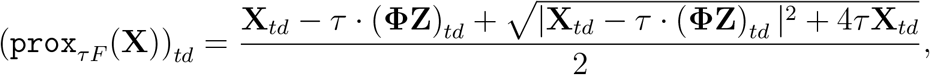

Where *τ* > 0. We further consider *G*_*m*_ (.) = ∥. ∥_1,_ *m* = ^1, 2^, and *K*_1_(R) := λ_time_D_2_R, *K*_2_(R) := λ_space_RB^*T*^ The proximity operators associated to *G*_*m*_ read:

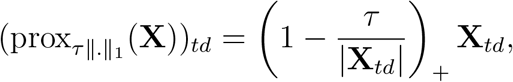

where (.)_+_ = max(0,.). In Algorithm 1, we express explicitly Algorithm (161) of [10] for our setting, considering the Moreau identity that provides the relation between the proximity operator of a function and the proximity operator of its conjugate (cf. Eq. (8) of [10]). The choice of the parameters *τ* and *σ*_*m*_ impacts the convergence guarantees. In this work, we adapt a standard choice provided by [5] to this extended framework. The adjoint of *K*_*m*_, denoted 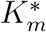, is given by 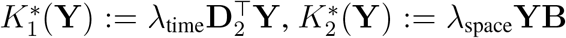. The sequence (**R**^(*k*+1)^)_*k∈*ℕ_ converges to a minimizer of (7) (cf. Thm 8.2 of [10]).

#### **Algorithm 1:** Chambolle-Pock with multiple penalization terms

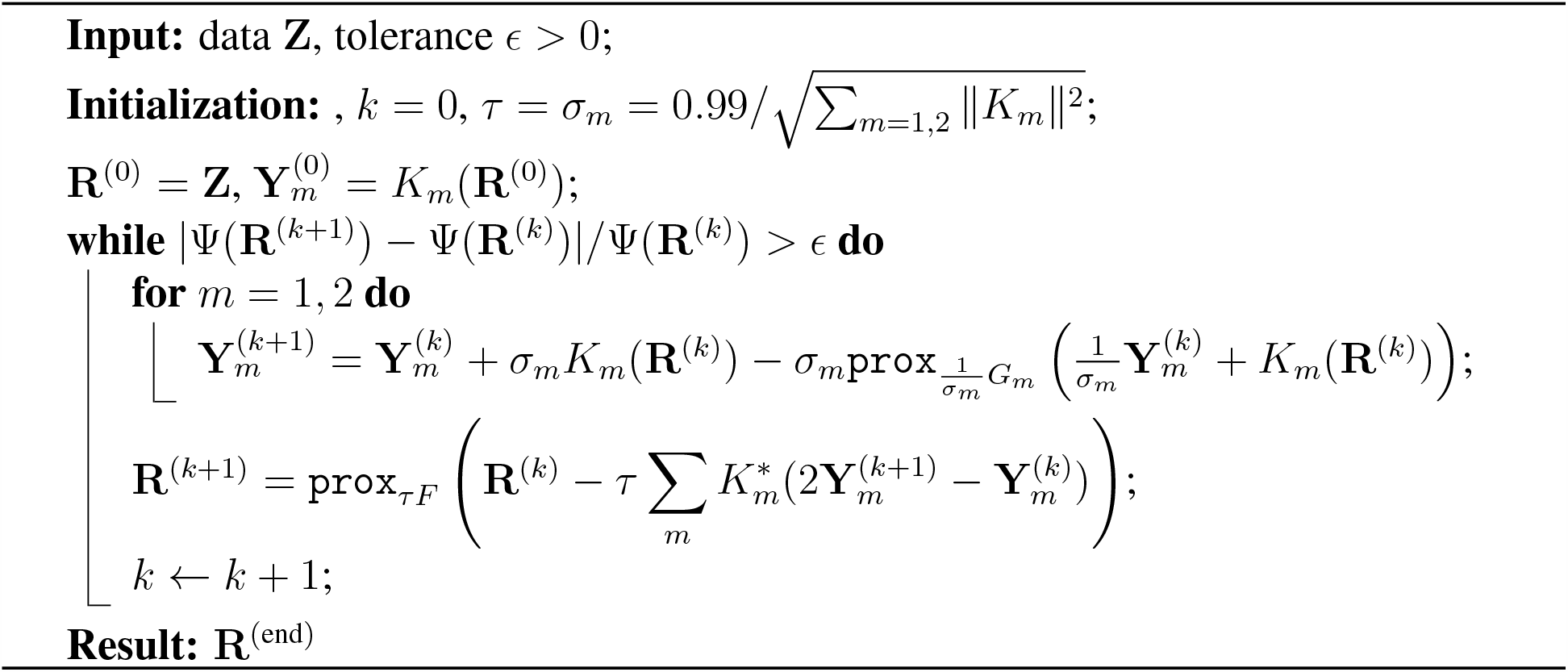

### 3.4 Estimation on synthetic data

To assess the relevance and performance of the proposed estimation procedure detailed above, it is first applied to two different synthetic time series *z*(*t*). The first one is synthesized using directly the model in Eq. (2), with the same serial interval function Φ(*t*) as that used for the estimation, and using an a priori prescribed function *R*(*t*). The second one is produced from solving a compartmental (SIR type) model. For such models, *R*(*t*) can be theoretically related to the time scale parameters entering their definition, as the ratio between the infection time scale and the quitting infection (be it by death or recovery) time scale. The theoretical serial function Φ associated to that model and to its parameters is computed analytically (cf., e.g., [6]) and used in the estimation procedure.

For both cases, the same a priori prescribed function *R*(*t*), to be estimated, is chosen as constant (*R* = 2.2) over the first 45 days to model the epidemic outbreak, followed by a linear decrease (till below 1) over the next 45 days to model lockdown benefits, and finally an abrupt linear increase for the last 10 days, modeling a possible outbreak at when lockdown is lifted. Additive Gaussian noise is superimposed to the data produced by the models to account for outliers and misreporting.

For both cases, the proposed estimation procedure (obtained with *λ*_time_ set to the same values as those used to analyze real data in Section 4) outperforms the naive estimates (1), which turn out to be very irregular (cf. Fig. 2). The proposed estimates notably capture well the three different phases of *R*(*t*) (stable, decreasing and increasing, with notably a rapid and accurate reaction to the increasing change in the 10 last days.

**Figure 2.**
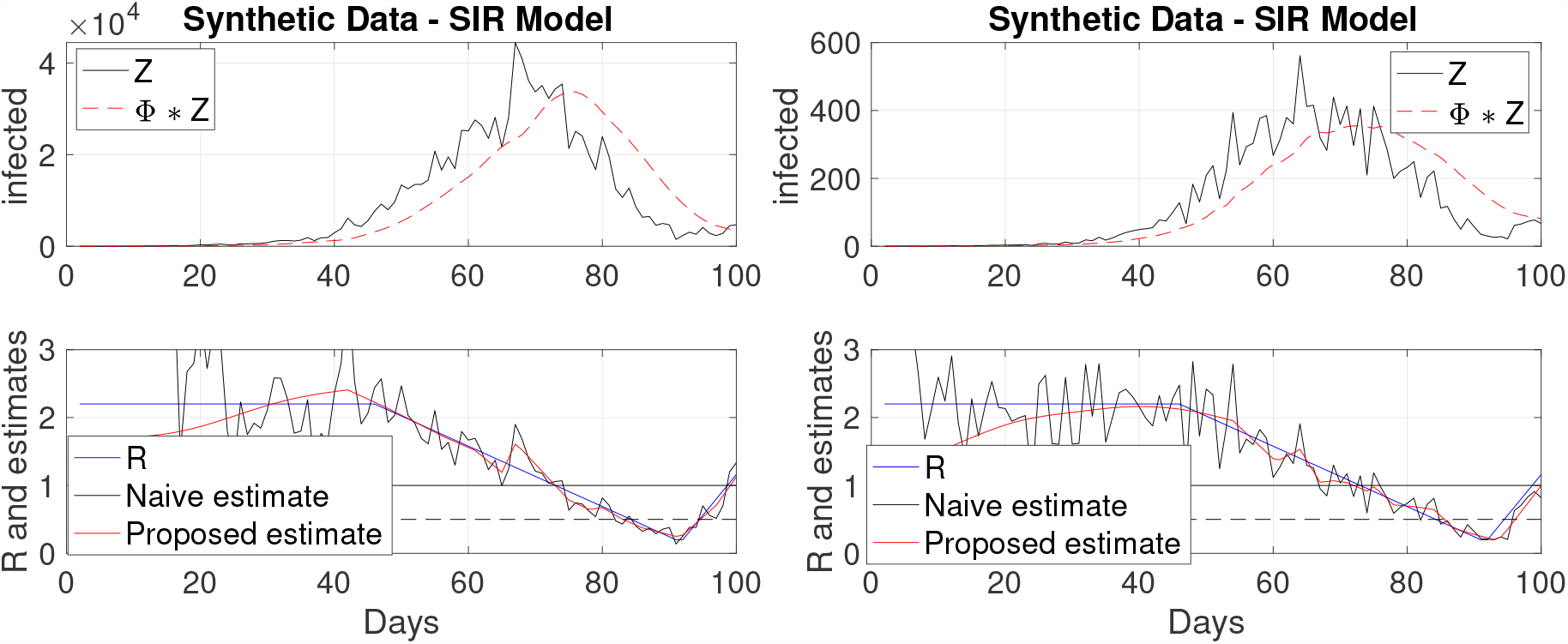
Estimated reproduction numbers *R*(*t*) on synthetic data, produced by the Poisson model (2) (left) and by a SIR model (right). The true *R*(*t*) (blue line) is piecewise linear: constant till day 45, decreasing till day 90 and increasing for the last 10 days. The proposed estimate (red) performs better than the naive estimate (black) (cf. Eq. (1)) and detects well the changes, notably it quickly reacts to the increase of the last 10 days.

## 4 COVID-19 reproduction number time evolutions

The present section aims to apply the models and estimation tools proposed in Section 3 to the data described in Section 2. First, methodological issues are addressed related to tuning the hyperparameter(s) *λ*_time_ or (*λ*_time_, *λ*_space_) in univariate and multivariate settings, and to comparing the consistency between different estimates of *R* obtained from the same incidence data yet downloaded from different repositories. Then, the estimation tools are applied to the estimation of *R*(*t*) independently for numerous countries (cf. Section 4.3) and jointly for the 94 continental France *départements* (cf. Section 4.4).

### 4.1 Regularization hyperparameter tuning

A critical issue associated with the practical use of the estimates based on the optimization problems (5) and (7) lies in the tuning of the hyperparameters balancing data fidelity terms and penalization terms. While automated and data-driven procedures can be devised, following works such as [20] and references therein, let us analyze the forms of the functional to be minimized, so as to compute relevant orders of magnitude for these hyperparameters.

Let us start with the univariate estimation (5). Using *λ*_time_ = 0 implies no regularization and the achieved estimate turns out to be as *noisy* as the one obtained with a naive estimator (cf. Eq. (1)). Conversely, for large enough *λ*_time_, the proposed estimate becomes *exactly* a constant, missing any time evolution. Tuning *λ*_time_ is thus critical but can become tedious, especially because differences across countries (or across *départements* in France) are likely to require different choices for *λ*_time_. However, a careful analysis of the functional to minimize shows that the data fidelity term (9), based on a Kullback-Leibler divergence, scales proportionally to the input incidence data *z* while the penalization term, based on the regularization of *R*(*t*), is independent of the actual values of *z*. Therefore, the same estimate for *R*(*t*) is obtained if we replace *z* with *α* × *z* and *λ* with *α* × *λ*. Because orders of magnitude of *z* are different amongst countries (either because of differences in population size, or of pandemic impact), this critical observation leads us to apply the estimate not to the raw data *z* but to a normalized version *z/*std(*z*), alleviating the burden of selecting one *λ*_time_ per country, instead enabling to select one same *λ*_time_ for all countries and further permitting to compare the estimated *R*(*t*)’s across countries for equivalent levels of regularization.

Considering now the graph-based spatially-regularized estimates (7) while keeping fixed *λ*_time_, the different *R*(*t*) are analyzed independently for each *département* when *λ*_space_ = 0. Conversely, choosing a large enough *λ*_space_ yields *exactly identical* estimates across *départments* that are, satisfactorily, very close to what is obtained from data aggregated over France prior to estimation. Further, the connectivity graph amongst the 94 continental France *départements* leads to an adjacency matrix with 475 non-zero off-diagonal entries (set to the value 1, associated to as many edges in the graph). Therefore, a careful examination of (7) shows that the spatial and temporal regularizations have equivalent weights when *λ*_time_ and *λ*_time_ are chosen such that

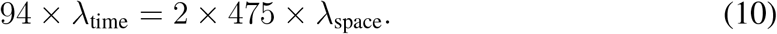

The use of *z/*std(*z*) and of (10) above gives a relevant first-order guess to the tuning of *λ*_time_ and of (*λ*_time_, *λ*_space_).

### 4.2 Estimate consistency using different repository sources

When undertaking such work dedicated to on-going events, to daily evolutions, and to a real stake in forecasting future trends, a solid access to reliable data is critical. As mentioned in Section 2, three sources of data are used, each including data for France, which are thus now used to assess the impact of data sources on estimated *R*(*t*). Source1(JHU) and Source2(ECDPC) provide cumulated numbers of confirmed cases counted at national levels and (in principle) including all reported cases from any source (hospital, death at home or in care homes…). Source3(SPF) does not report that same number, but a collection of other figures related to hospital counts only, from which a daily number of new hospitalizations can be reconstructed and used as a proxy for daily new infections. The corresponding raw and (sliding-median) preprocessed data, illustrated in Fig. 3, show overall comparable shapes and evolutions, yet with clearly visible discrepancies of two kinds. First, Source1(JHU) and Source2(ECDPC), consisting of crude reports of number of confirmed cases are prone to outliers. Those can result from miscounts, from pointwise incorporations of new figures, such as the progressive inclusion of cases from *EHPAD* (care homes) in France, or from corrections of previous erroneous reports. Conversely, data from Source3(SPF), based on hospital reports, suffer from far less outliers, yet at the cost of providing only partial figures. As discussed in Section 2.2, it has been chosen here to preprocess outliers, on the basis of a sliding median procedure, prior to conducting the estimation of *R*(*t*). Second, in France, as in numerous other countries worldwide, the procedure on which confirmed case counts are based, changed several times during the pandemic period, yielding possibly some artificial increase in the local average number of daily new confirmed cases. This has notably been the case for France, prior to the end of the lockdown period (mid-May), when the number of tests performed has regularly increased for about two weeks, or more recently early June when the count procedures has been changed again, likely because of the massive use of serology tests. Because the estimate of *R*(*t*) essentially relies on comparing a daily number against a past moving average, these changes lead to significant biases that cannot be easily accounted for, but vanishes after some duration controlled by the typical width of the serial distribution Φ (of the order of ten days).

**Figure 3.**
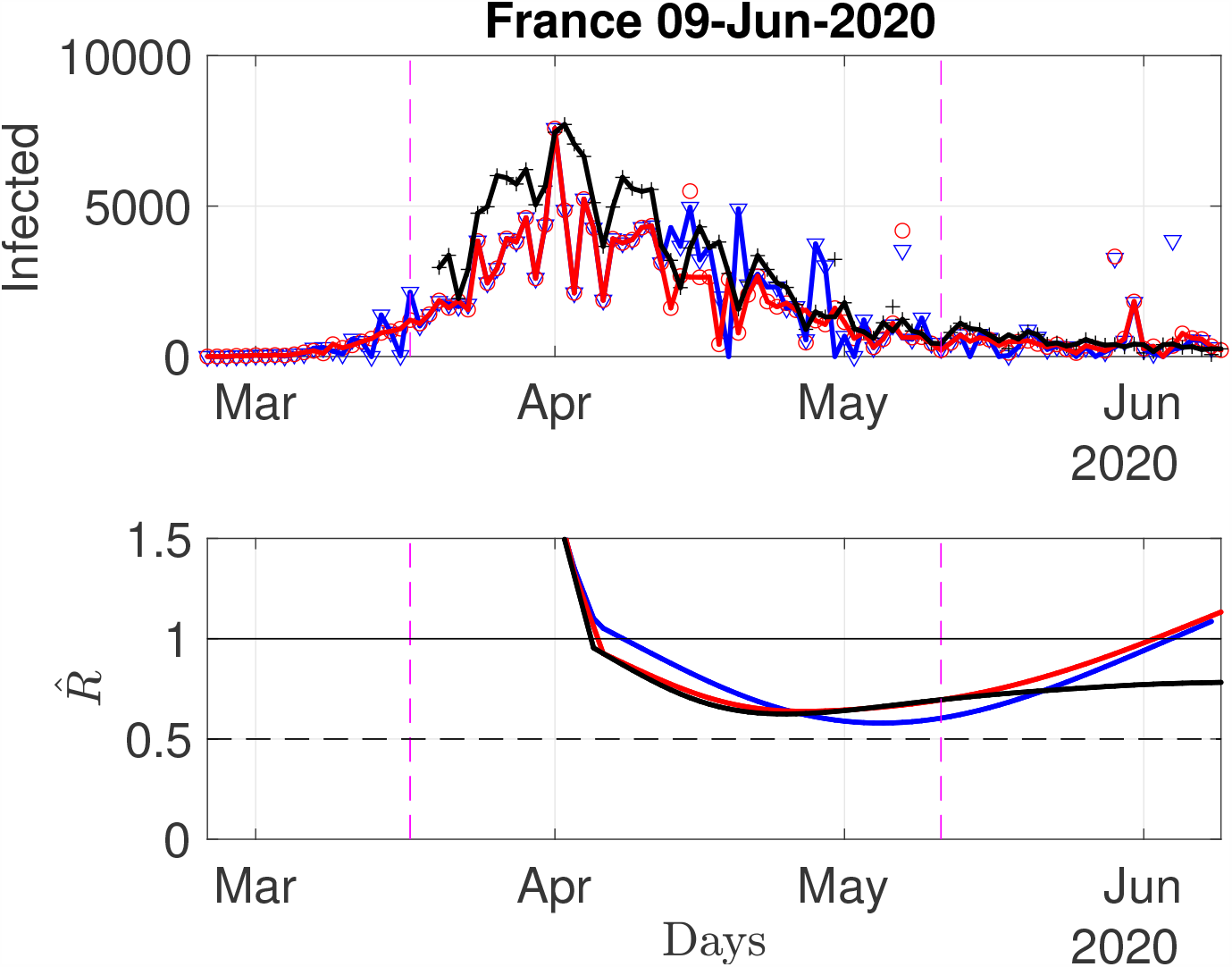
Daily new confirmed cases for France, from three different sources. Top row: raw data (symbols) and sliding median preprocessed data (connected lines) from Source1(JHU) (blue) and Source2(ECDPC)(red) and Source3(SPF) (black). Bottom row: corresponding estimates of *R*(*t*).

Fig. 3 further compares, for a relevant value of *λ*_time_ selected as explicitly discussed below in Section 4.3, estimates obtained from the three different sources of data. Overall shapes in the time evolution of estimates are consistent, For instance, the three sources led to estimates showing a mild yet clear increase of *R*(*t*) for the period ranging from early May to May 20th, likely corresponding to a bias induced by the regular increase of tests actually performed in France. These comparisons however also clearly show that estimates are impacted by outliers and thus do depend on preprocessing. These considerations led to the final choice, used hereafter, of a threshold of ±2.5 std, in the sliding median denoising of Section 2.2.

### 4.3 Confirmed infection cases across the world

To report estimated *R*(*t*)’s for different countries, data from Source2(ECDPC) are used as they are of better quality than data from Source1(JHU), and because hospital-based data (as in Source3(SPF)) are not easily available for numerous different countries. Visual inspection led us to choose, uniformly for all countries, two values of the temporal regularization parameter: *λ*_time_ = 50 to produce a strongly-regularized, hence slowly varying estimate, and *λ*_time_ = 3.5 for a milder regularization, and hence a more reactive estimate. These estimates being by construction designed to favor piecewise linear behaviors, local trends can be estimated by computing (robust) estimates of the derivatives 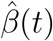 Of 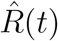. The slow and less slow estimates of 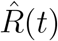 thus provide a slow and less slow estimate of the local trends. Intuitively, these local trends can be seen as predictors for the forthcoming value of 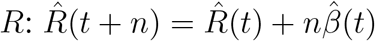.

Let us start by inspecting again data for France, further comparing estimates stemming from data in Source2(ECDPC) or in Source3(SPF) (cf. Fig. 4). As discussed earlier, data from Source2(ECDPC) show far more outliers that data from Source3(SPF), thus impacting estimation of *R* and *β*. As expected, the strongly regularized estimates (*λ*_time_ = 50) are less sensitive than the less regularized ones (*λ*_time_ = 3.5), yet discrepancies in estimates are significant, as data from Source2(ECDPC) yields, for June 9th, estimates of *R* slightly above 1, while that from Source3(SPF) remain steadily around 0.75, with no or mild local trends. Again, this might be because late May, France has started massive serology testing, mostly performed outside hospitals. This yielded an abrupt increase in the number of new confirmed cases, biasing upward the estimates of *R*(*t*). However, the short-term local trend for June 9th goes also downward, suggesting that the model is incorporating these irregularities and that estimates will return to unbiased after an estimation time controlled by the typical width of the serial distribution Φ (of the order of ten days). This recent increase is not seen in Source3(SPF)-based estimates that remain very stable, potentially suggesting that hospital-based data are much less sensitive to changes in the count policies.

**Figure 4.**
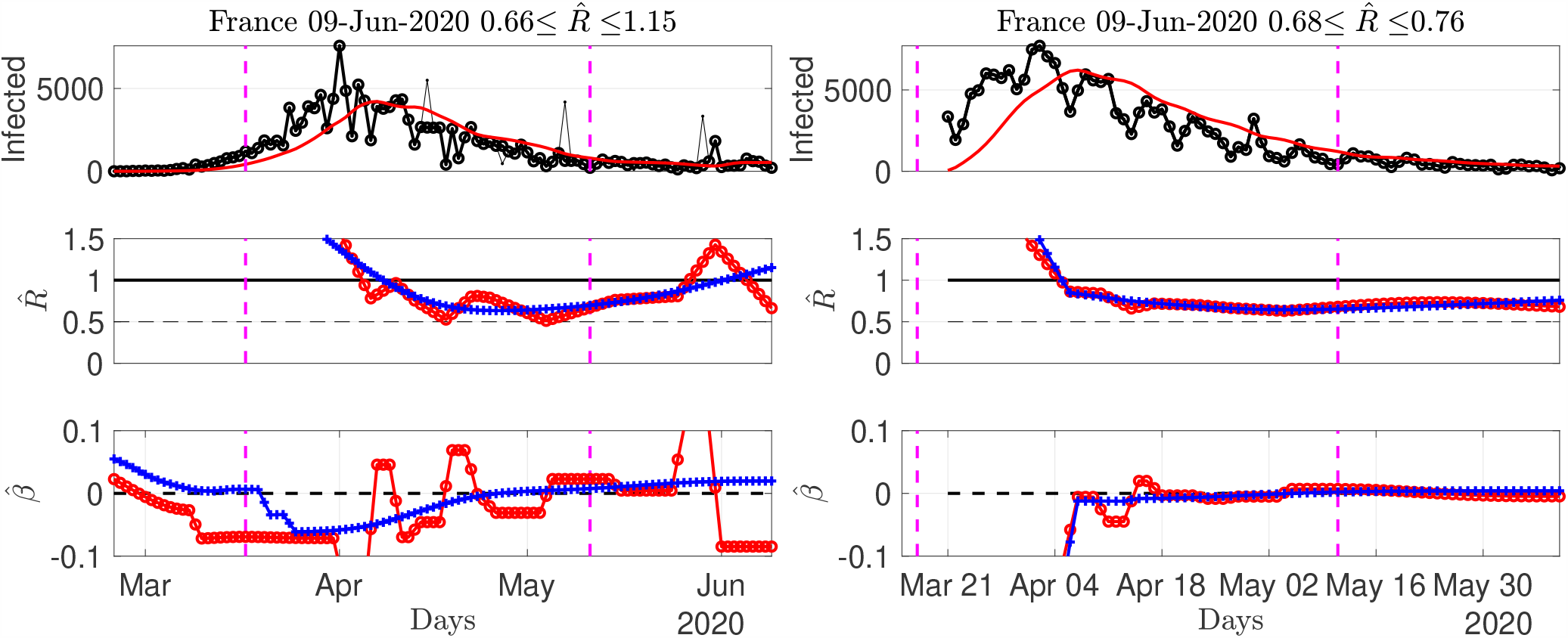
Number of daily new confirmed cases and reproduction number and local estimates for France, using data from Source2(ECDPC) (left) and Source3(SPF) (right) (reconstructed proxy from hospital counts). Top: time series. Middle: fast (red) and slowly evolving (blue) estimates of *R*(*t*). Bottom: fast (red) and slowly evolving (blue) estimates of local trends *β*(*t*). The title of the plots report estimates for the current day.

Source2(ECDPC) provides data for several tens of countries. Figs. 5 to 8 report 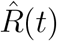 and 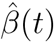 for several selected countries (more figures are available at perso.enslyon.fr/patrice.abry). As of June 9th (time of writing), Fig. 5 shows that for most European countries, the pandemic seems to remain under control despite lifting of the lockdown, with (slowly varying) estimates of *R* remaining stable below 1, ranging from 0.7 to 0.8 depending on countries, and (slowly varying) trends around 0. Sweden and Portugal (not shown here) display less favorable patterns, as well as, to a lesser extent, The Netherlands, raising the question of whether this might be a potential consequence of less stringent lockdown rules compared to neighboring European countries. Fig. 6 shows that while 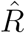 for Canada is clearly below 1 since early May, with a negative local trend, the USA are still bouncing back and forth around 1. South America is in the above 1 phase but starts to show negative local trends. Fig. 7 indicates that Iran, India or Indonesia are in the critical phase with 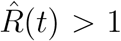. Fig. 8 shows that data for African countries are uneasy to analyze, and that several countries such as Egypt or South Africa are in pandemic growing phases.

**Figure 5.**
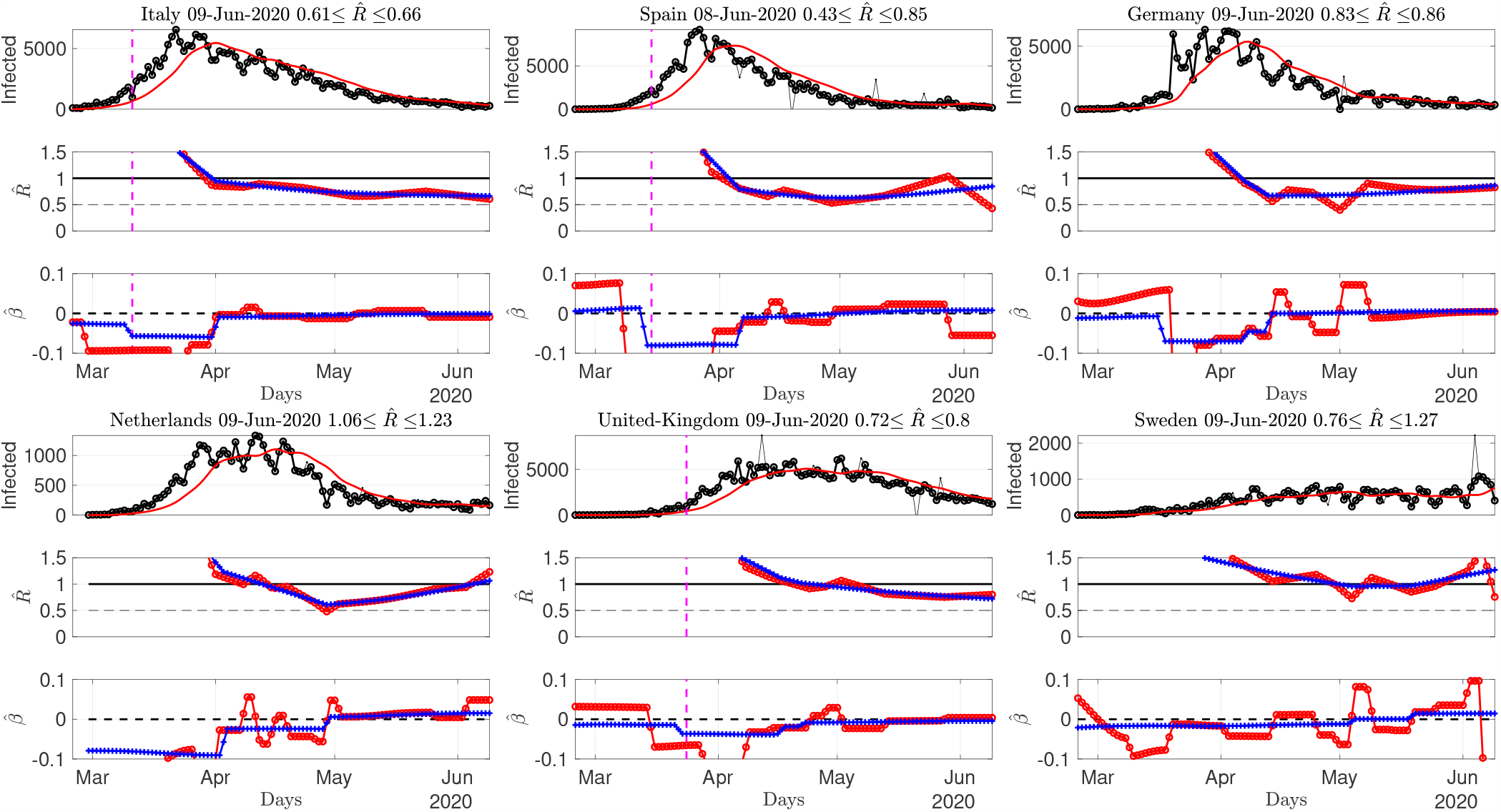
Number of daily new confirmed cases and reproduction number and local estimates for countries in Europe. Top: time series. Middle: fast (red) and slowly evolving (blue) estimates of *R*(*t*). Bottom: fast (red) and slowly evolving (blue) estimates of local trends *β*(*t*). The title of the plots report estimates for the current day. Data from Source2(ECDPC).

**Figure 6.**
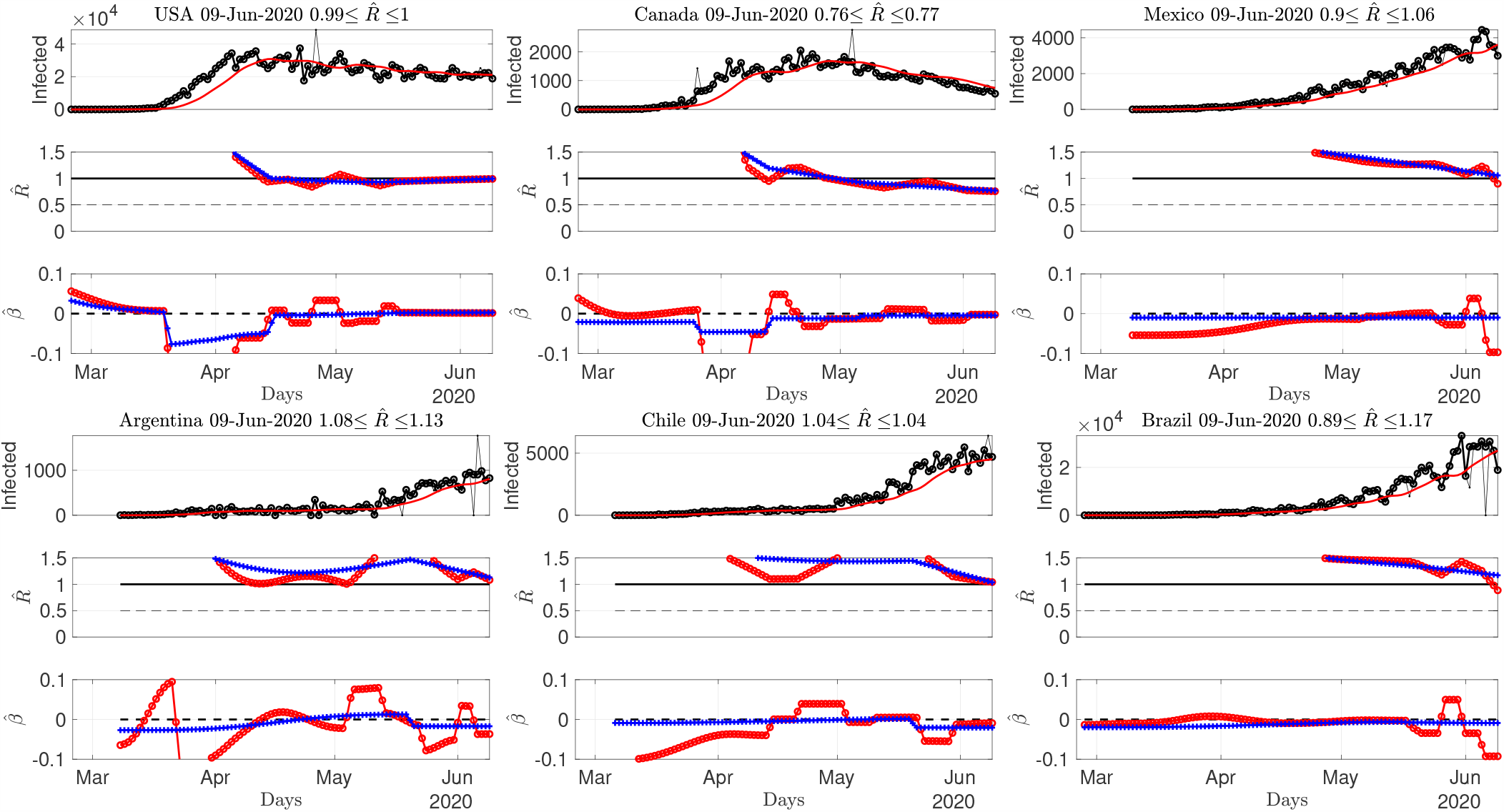
Number of daily new confirmed cases and reproduction number and local estimates for American countries. Top: time series. Middle: fast (red) and slowly evolving (blue) estimates of *R*(*t*). Bottom: fast (red) and slowly evolving (blue) estimates of local trends *β*(*t*). The title of the plots report estimates for the current day. Data from Source2(ECDPC).

**Figure 7.**
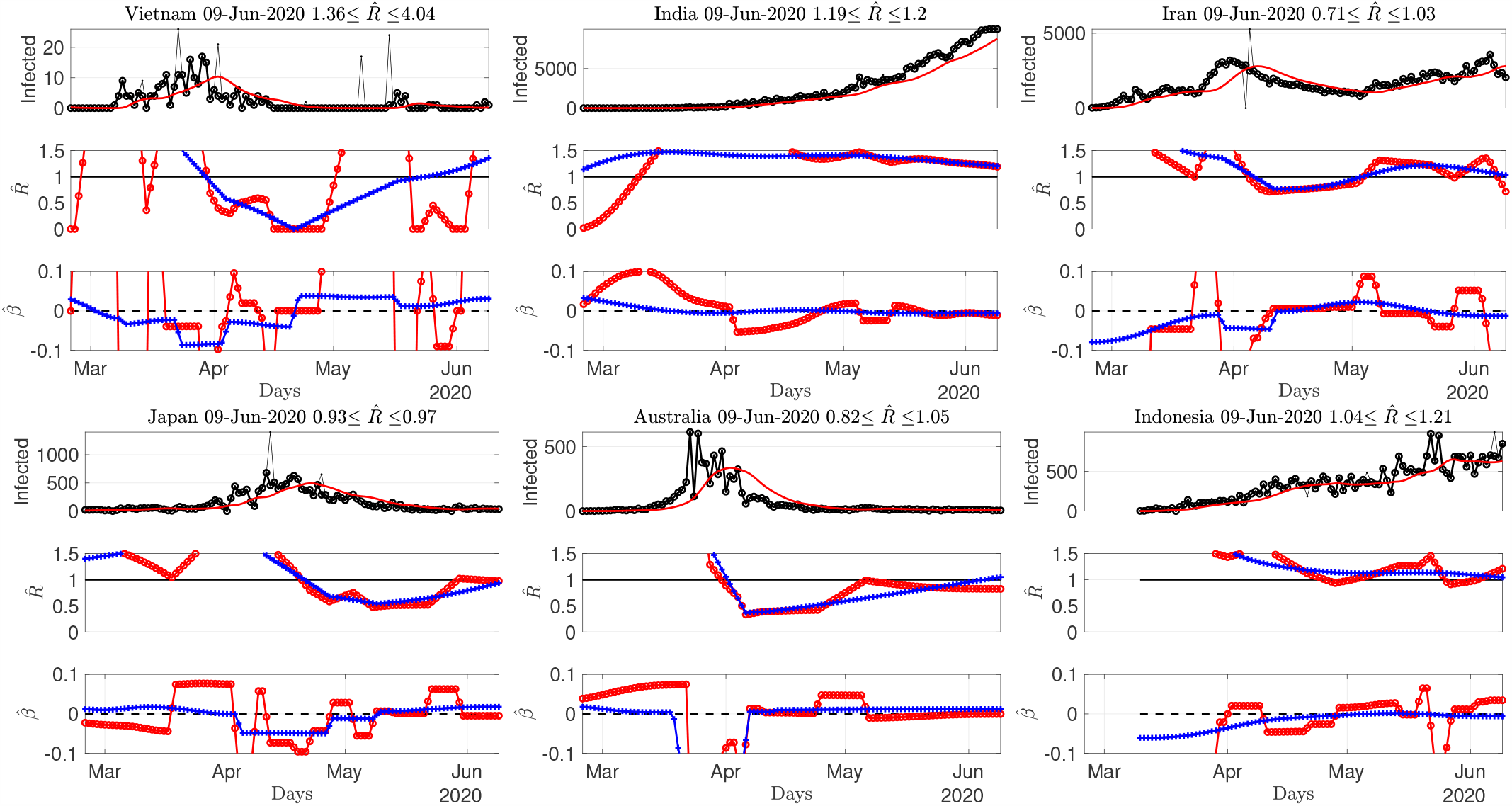
Number of daily new confirmed cases and reproduction number and local estimates for Asian countries. Top: time series. Middle: fast (red) and slowly evolving (blue) estimates of *R*(*t*). Bottom: fast (red) and slowly evolving (blue) estimates of local trends *β*(*t*). The title of the plots report estimates for the current day. Data from Source2(ECDPC).

**Figure 8.**
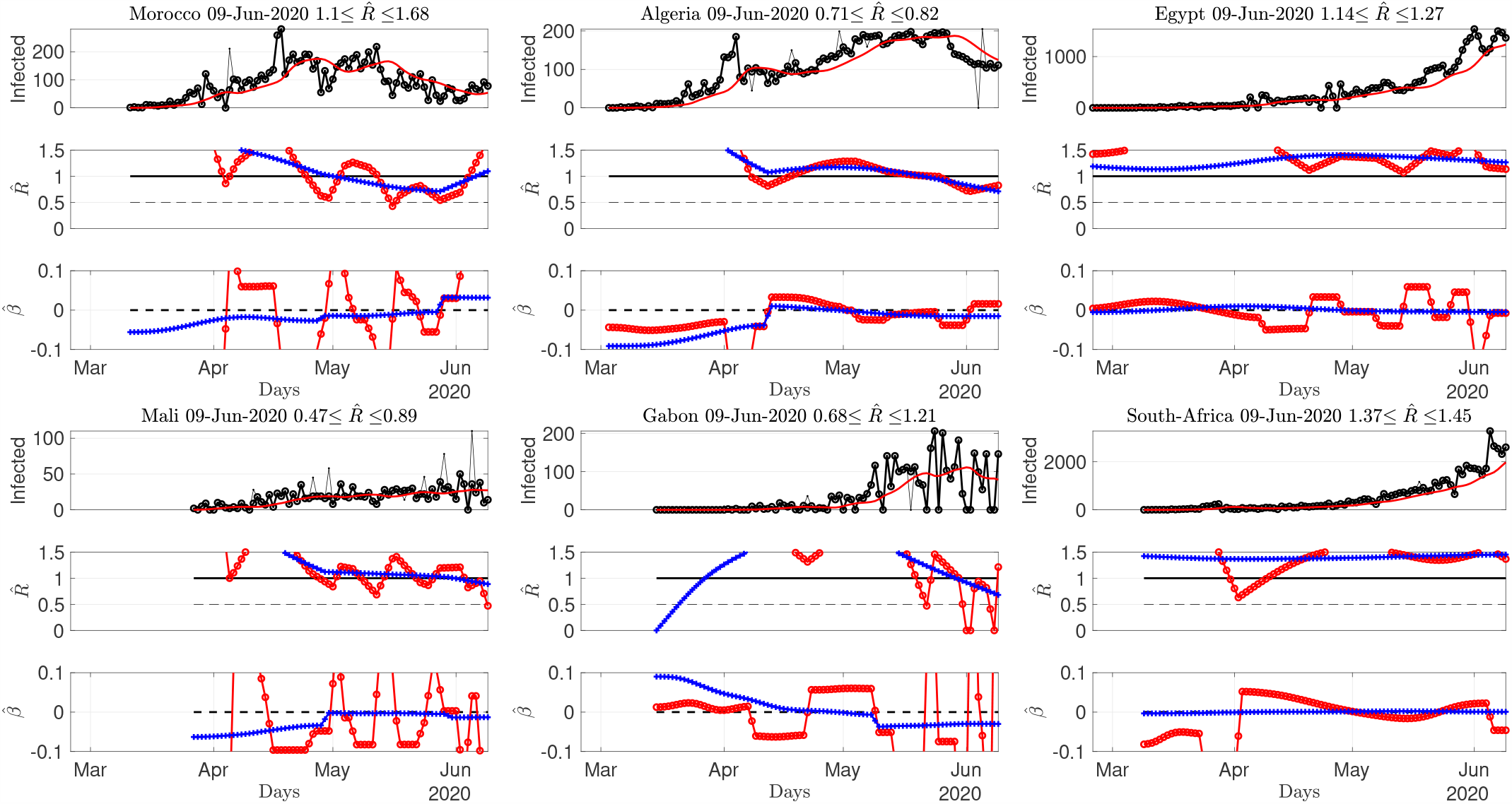
Number of daily new confirmed cases and reproduction number and local estimates for African countries. Top: time series. Middle: fast (red) and slowly evolving (blue) estimates of *R*(*t*). Bottom: fast (red) and slowly evolving (blue) estimates of local trends *β*(*t*). The title of the plots report estimates for the current day. Data from Source2(ECDPC).

#### Phase-space representation

To complement Figs. 5 to 8, Fig. 9 displays a phase-space representation of the time evolution of the pandemic, constructed by plotting one against the other local averages over a week of the slowly varying estimated reproduction number 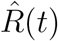 and local trend, 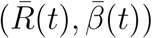, for a period ranging from mid-April to June 9th. Country names are written at the end (last day) of the trajectories. Interestingly, European countries display a C-shape trajectory, starting with *R >* 1 with negative trends (lockdown effects), thus reaching the safe zone (*R <* 1) but eventually performing a U-turn with a slow increase of local trends till positive. This results in a mild but clear re-increase of *R*, yet with most values below 1 today, except for France (see comments above) and Sweden. The USA display a similar C-shape though almost concentrated on the edge point *R*(*t*) = 1, *β* = 0, while Canada does return to the safe zone with a specific pattern. South-American countries, obviously at an earlier stage of the pandemic, show an inverted C-shape pattern, with trajectory evolving from the bad top right corner, to the *controlling phase* (negative local trend, with decreasing *R* still above 1 though). Phase-spaces of Asian and African countries essentially confirm these C-shaped trajectories. Envisioning these phase-space plots as pertaining to different stages of the pandemic (rather than to different countries), this suggests that COVID-19 pandemic trajectory ressembles a counter-clock wise circle, starting from the *bad* top right corner (*R* above 1 and positive trends), evolving, likely by lockdown impact, towards the bottom right corner (*R* still above 1 but negative trends) and finally to the *safe* bottom left corner (*R* below1 and negative then null trend). The lifting of the lockdown may explain the continuation of the trajectory in the *still safe but*… corner (*R* below1 and again positive trend). As of June 9th, it can be only expected that trajectories will not close the loop and reach back the *bad* top right corner and the *R* = 1 limit.

**Figure 9.**
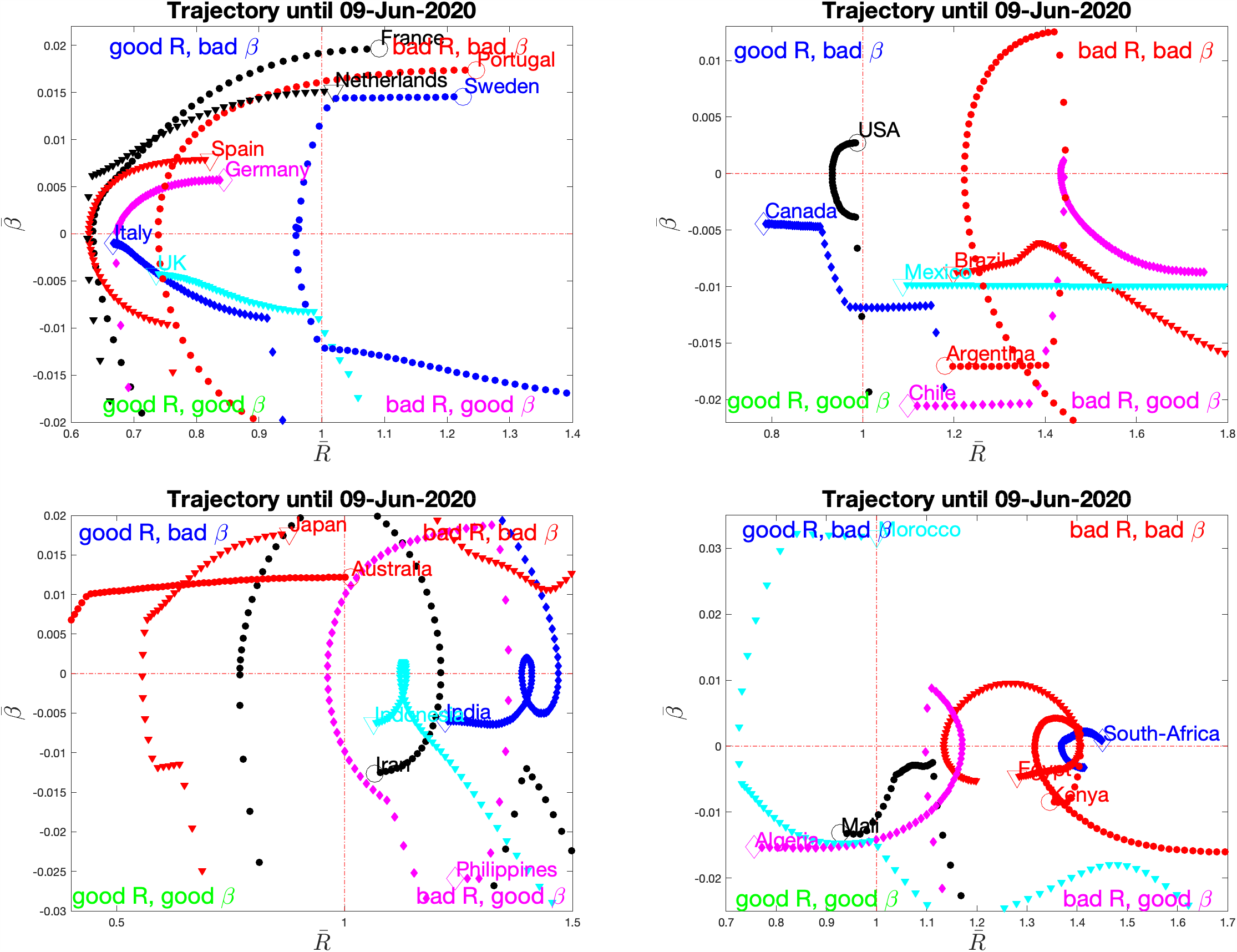
Phase-space evolution. reconstructed from averaged slowly varying estimates of *R* and *β*, per continent. The name of the country is written at the last day of the trajectory, also marked by larger size empty symbol. Data from Source2(ECDPC).

### 4.4 Continental France *départements*: regularized joint estimates

There is further interest in focusing the analysis on the potential heterogeneity in the epidemic propagation across a given territory, governed by the same sanitary rules and health care system. This can be achieved by estimating a set of *local* 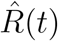’s for different provinces and regions [14]. Such a study is made possible by the data from Source3(SPF), that provides hospital-based data for each of the continental France *départements*. Fig. 4 (right) already reported the slow and fast varying estimates of *R* and local trends computed from data aggregated over the whole France. To further study the variability across the continental France territory, the graph-based, joint spatial and temporal regularization described in Eq. 7 (cf. Section 3.2) is applied to the number of confirmed cases consisting of a matrix of size *K × T*, with *D* = 94 continental France *départements*, and *T* the number of available daily data (e.g., *T* = 78 on June 9th data being available only after March, 18th). The two choices for *λ*_time_ leading to slow and less slow estimates were kept for this joint study. Using (10) as a guideline, empirical analyses led to set *λ*_space_ = 0.025, thus selecting spatial regularization to weight one-fourth of the temporal regularization.

First, Fig. 10 (top row) maps and compares for June 9th (chosen arbitrarily as the day of writing) per-*département* estimates, obtained when *départements* are analyzed either independently (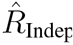 using Eq. 6, left plot) or jointly (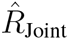 using Eq. 7, right plot). While the means of 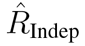 and 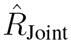 are of the same order (≃ 0.58 and ≃ 0.63 respectively) the standard deviations drop down from ≃ 0.40 to ≃ 0.14, thus indicating a significant decrease in the variability across departments. This is further complemented by the visual inspection of the maps which reveals reduced discrepancies across neighboring departments, as induced by the estimation procedure.

**Figure 10.**
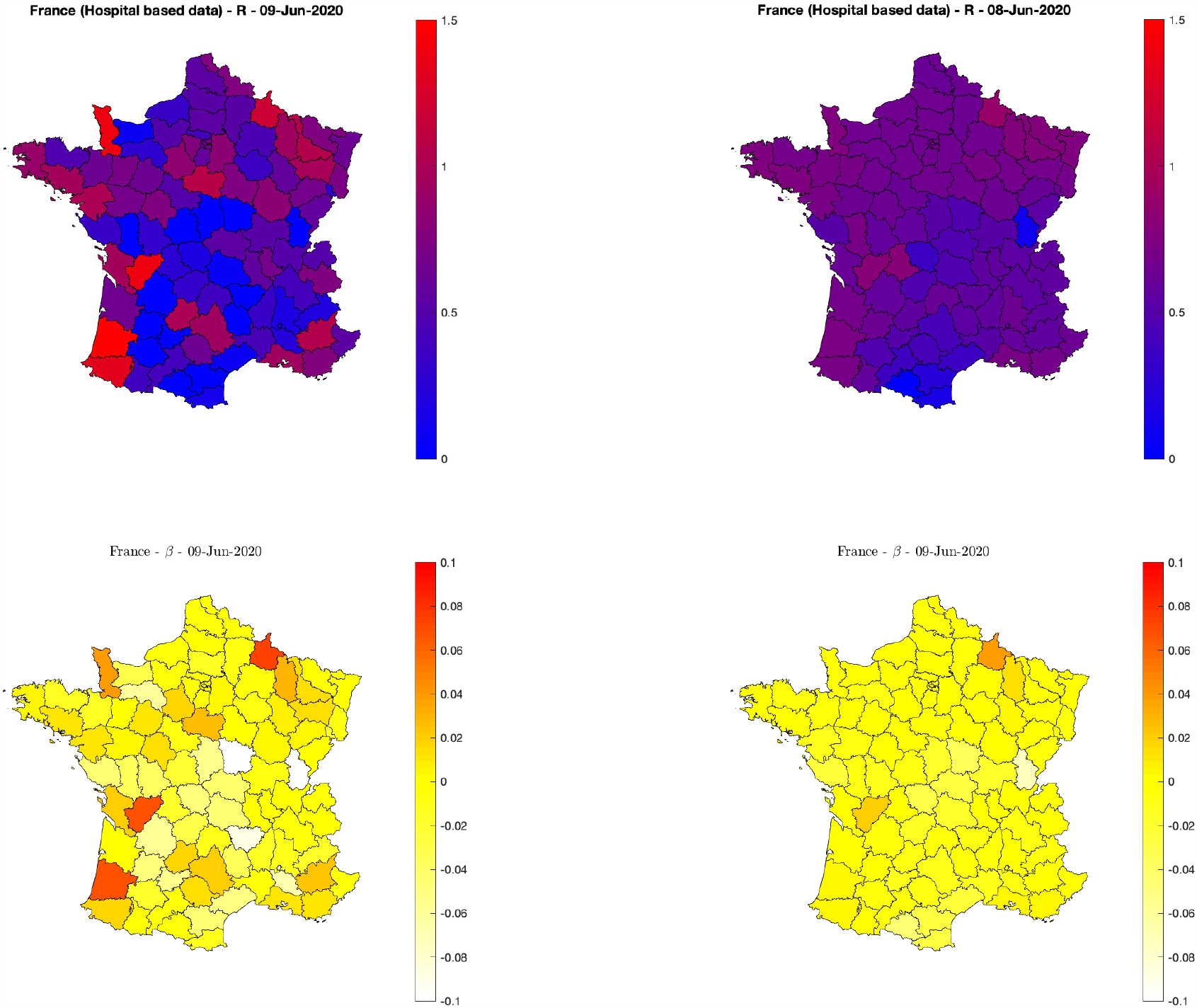
Reproduction numbers and trends for continental France *départements*. Fast varying estimates of reproduction numbers *R* (top) and trends *β* (bottom) for independent (left) and spatial graph-based regularized estimates (right). Hospital-based data from Source3(SPF).

In a second step, short and long-term trends are automatically extracted from 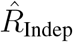 and 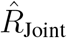 and short-term trends are displayed in the bottom row of Fig. 10 (left and right, respectively). This evidences again a reduced variability across neighboring departments, though much less than that observed for 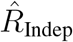 and 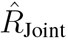, likely suggesting that trends on *R* per se are more robust quantities to estimate than single *R*’s. For June 9th, Fig. 10 also indicates globally mild decreasing trends (−0.007 ±0.010 per day, on average) everywhere across France, thus confirming the trend estimated on data aggregated over all France (cf. Fig. 4, right plot).

A video animation, available at perso.ens-lyon.fr/patrice.abry/DeptRegul.mp4 and updated on a daily basis (see also barthes.enssib.fr/coronavirus/IXXI-SiSyPhe/), reports further comparisons between 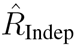 and 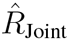 and their evolution along time for the whole period of data availability. Maps for selected days are also displayed in Fig. 11 (with identical colormaps and colorbars across time). Fig. 11 shows that until late March (lockdown took place in France on March, 17th), 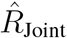 was uniformly above 1.5 (chosen as the upper limit of the colorbar to permit to see variations during the lockdown and post-lockdown periods), indicating a rapid evolution of the epidemic across entire France. A slowdown of the epidemic evolution is visible as early as the first days of April (with overall decreases of 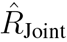, and a clear North vs. South gradient). During April, this gradient rotates slightly and aligns on a North-East vs. South-West direction and globally decreases in amplitude. Interestingly, in May, this gradient has reversed direction from South-West to North-East, though with very mild amplitude.

**Figure 11.**
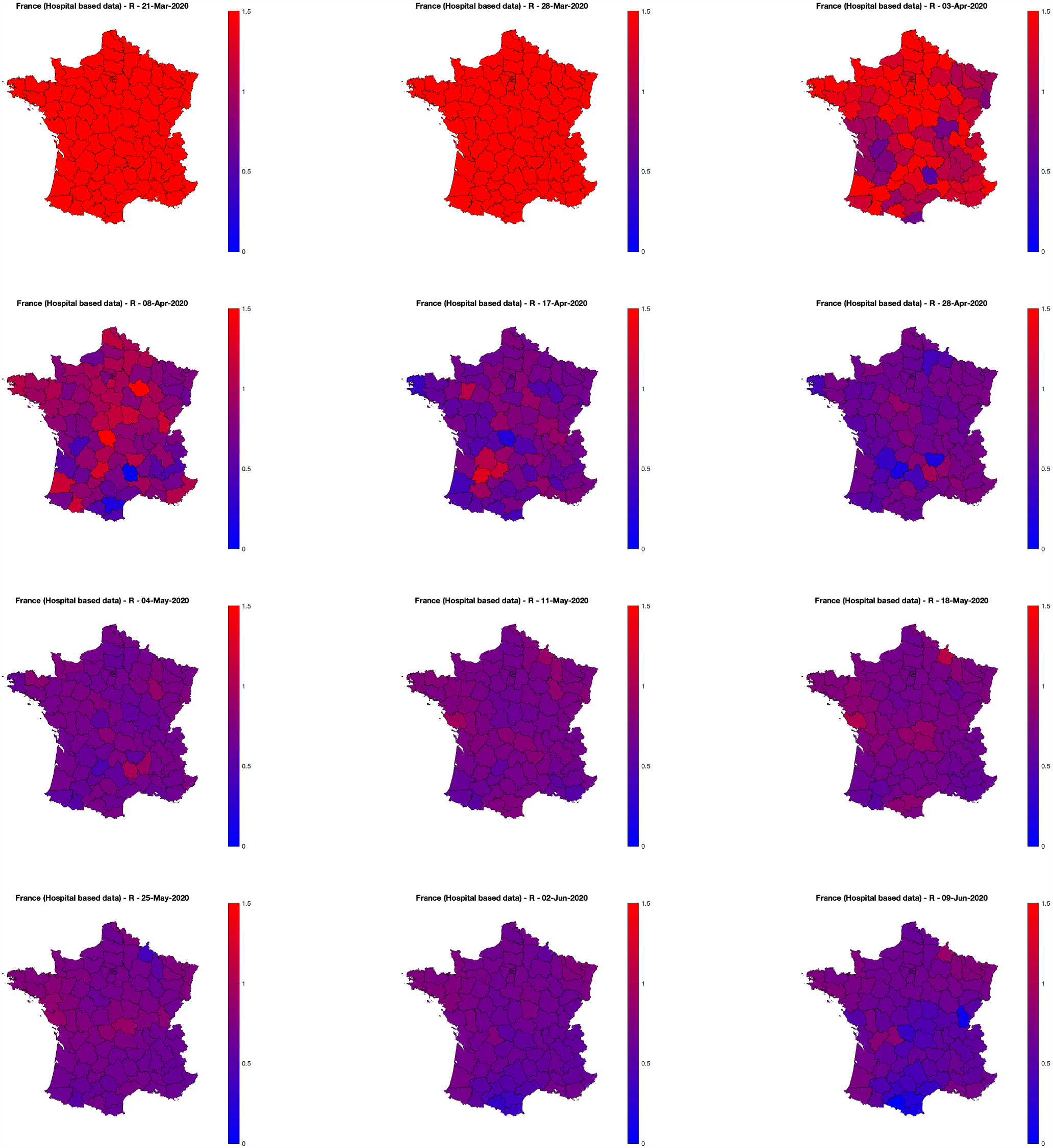
Graph-based spatially regularized estimates of the reproduction number *R* for the 94 continental France *départments*, as a function of days. Movie animations for the whole period are made available at perso.ens-lyon.fr/patrice.abry/DeptRegul.mp4 or barthes.enssib.fr/coronavirus/IXXI-SiSyPhe/, and updated on a regular basis. Hospital-based data from Source3 (SPF).

As of today (June 9th), the pandemic, viewed Hospital-based data from Source3(SPF), seems under control for the whole continental France.

## 5 Conclusions and perspectives

The estimation of reproduction numbers constitutes a classical task in assessing the status of a pandemic. Classically, this is done a posteriori (after the pandemic) and from consolidated data, often relying on detailed and accurate SIR-based models and making use of Bayesian frameworks. However, on-the-fly monitoring of the reproduction number time evolution constitutes a critical societal stake in situations such as that of COVID-19, when decisions need to be taken and action need to be made under emergency. This calls for a triplet of constraints: i) robust access to fast-collected data; ii) semi-parametric models for such data that focus on a subset of critical parameters; iii) estimation procedures that are elaborated enough to yield robust estimates and versatile enough to be used in quasi-real time (daily basis) and applied to (often-limited in quality and quantity) available data. In that spirit, making use of a robust nonstationary Poisson-distribution based semi-parametric model proven robust in the literature for epidemic analysis, we developed an original estimation procedure to favor piecewise regular estimation of the evolution of the reproduction number, both along time and across space. This was constructed as an inverse problem formulation designed to achieve robustness in the estimation by enforcing time and space smoothness through regularization while permitting fast enough temporal and spatial evolutions and solved using used proximal operators and nonsmooth convex optimization. The proposed tools were applied to pandemic incidence data consisting of daily counts of new infections, from several databases providing data either worldwide on an aggregated per-country basis or, for France only, based on the sole hospital counts, spread across the French territory. They permitted to reveal interesting patterns on the state of the pandemic across the world as well as to assess variability across one single territory governed by the same (health care and politics) rules.

At the practical level, this tool can be applied to time series of incidence data, reported, e.g., for a given country. Whenever made possible from data, estimation can benefit from a graph of spatial proximity between subdivisions of a given territory. Importantly, this tool can be used everyday easily as an on-the-fly monitoring procedure for assessing the current state of the pandemic and predict its short-term future evolution. Indeed, the tool also provides local trends *β*, whose last value can be used to forecast short-term future values of *R* and thus to detect a sudden increase of the pandemic.

The tools can be made available upon motivated request. Achieved estimations are updated on a daily basis at perso.ens-lyon.fr/patrice.abry (see also barthes.enssib.fr/coronavirus/IXXI-SiSyPhe/).

At the methodological level, the tool can be further improved in several ways. Instead of using Ω(**R**) := *λ*_time_ ∥ **D**_2_**R ∥**_1_ + *λ*_space_ ∥**RB**^*T*^∥ _1_, for the joint time and space regularization, another possible choice is to directly consider the matrix **D**_2_**RB**^*T*^ of joint spatiotemporal derivatives, and to promote sparsity with an */J*_1_-norm, or structured sparsity with a mixed norm *ℓ*_1,2_, e.g., ∥**D**_2_**RB**^*T*^*∥*_1,2_ = ∑_*t*_ ∥ (**D**_2_**RB**^*T*^)(*t*) ∥_2_. As previously discussed, data collected in the process of a pandemic are prone to several causes for outliers. Here, outlier preprocessing and reproduction number estimation were conducted in two independent steps, which can turn suboptimal. They can be combined into a single step at the cost of increasing the representation space permitting to split observation in true data and outliers, by adding to the functional to minimize an extra regularization term and devising the corresponding optimization procedure, which becomes nonconvex, and hence far more complicated to address. Finally, when an epidemic model suggests a way to make use of several time series (such as, e.g., infected and deceased) for one same territory, the tool can straightforwardly be extended into a multivariate setting by a mild adaptation of optimization problems (6) and (7), replacing the Kullback-Leibler divergence *D*_KL_(**Z** | **R** ⊙ **ΦZ**) by 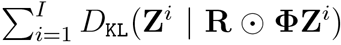. Finally, automating a data-driven tuning of the regularization hyperparameters constitutes another important research track.

## Data Availability

All the data processed in this paper is publicly available in repositories.

https://raw.githubusercontent.com/CSSEGISandData/COVID-19/master/csse_covid_19_time_series/

https://www.ecdc.europa.eu/sites/default/files/documents/COVID-19-geographic-disbtribution-worldwide.xlsx

https://coronavirus.jhu.edu/

https://www.ecdc.europa.eu/

https://www.santepubliquefrance.fr/

## Footnotes

1 https://coronavirus.jhu.edu/ and https://raw.githubusercontent.com/CSSEGISandData/COVID-19/master/csse_covid_19_time_series/

2 https://www.ecdc.europa.eu/ and https://www.ecdc.europa.eu/sites/default/files/documents/COVID-19-geographic-disbtribution-worldwide.xlsx

3 https://www.santepubliquefrance.fr/ and https://www.data.gouv.fr/fr/datasets/

